# “I walk around like my hands are covered in mud”: food safety and hand hygiene behaviours of Canadians during the COVID-19 pandemic

**DOI:** 10.1101/2020.08.25.20181545

**Authors:** Robyn Haas, Fatih Sekercioglu, Richard Meldrum, Ian Young

## Abstract

**Objectives:** To investigate how and why Canadians engaged in different food handling and hand hygiene behaviours during the COVID-19 pandemic.

**Methods:** Seven online, text-based focus groups were conducted with a total of 42 participants. Eligible participants included adults living in Canada that prepared meals at home at least once per week. Focus groups took place from May-June 2020 and followed a semi-structured question guide. Participants were asked about their practices relating to food preparation habits at home; food purchasing, handling, and storage; hand hygiene and sanitation; and information sources about food safety concerning the COVID-19 pandemic. A thematic analysis was conducted using the Theoretical Domains Framework as a coding guide.

**Results:** The most notable changes in behaviour since the onset of the pandemic were seen in participants’ handwashing, sanitation, and grocery shopping practices. Participants tended to perceive grocery store employees, shoppers, and food service staff as having inadequate sanitation precautions and, therefore, as a source of COVID- 19 transmission risk. They heavily relied on public health, medical, and government officials as sources of information. Feelings of stress and anxiety appeared to be linked to certain sanitation behaviours. Many participants displayed a general apathy toward routine food safety practices such as safe food storage at home.

**Conclusion:** This work supports the need for clear and concise messaging for hand hygiene and food safety behaviours during the COVID-19 pandemic and in future times of crisis. It also highlights a need for ongoing food safety messaging.

## Introduction

Foodborne illness has a significant burden on morbidity and mortality in Canada. It is estimated that 4 million Canadians become ill due to foodborne illness annually, leading to more than 11,000 hospitalizations and over 200 deaths (Thomas et al., 2015). Furthermore, most cases of foodborne illness go undetected and are, therefore underreported (MacDougall et al., 2008). Foodborne illnesses result in a multitude of economic impacts leading to significant costs to the healthcare system (i.e. physician time, lab tests) and society due to lost productivity. Furthermore, their associated burden on the healthcare system is exacerbated during times of crisis when the system is already at or approaching capacity, such as the coronavirus disease 2019 (COVID-19) pandemic (Bialek et al., 2020). Previous Canadian studies have shown that consumers often do not engage in recommended safe food handling practices and behaviours at home, which can increase their risk of foodborne illness (Murray et al., 2017; Nesbitt et al., 2014). This is particularly noteworthy during the COVID-19 pandemic as many restaurants were closed during its early stage, resulting in more cooking at home. As such, additional efforts are needed by consumers during pandemic situations to prevent foodborne illness and its severe consequences in order to help relieve stress on the healthcare system.

At the onset of the COVID-19 pandemic, media reports noted a trend among consumers of “panic buying” or stockpiling a variety of household supplies and foods, including perishable items such a meat and poultry (Wiener-Bronner, 2020). This trend was also noted during prior emergencies, such as a 2015 snowstorm in New York City (Zheng, Shou, & Yang, 2020). This practice raises food safety concerns due to the need for adequate storage and refrigeration of these foods, which must either be frozen or consumed within three days to prevent hazardous microbial growth (Health Canada, 2019). Additionally, hand sanitizer shortages led to online promotion of homemade hand sanitizer recipes, which have limitations compared to traditional handwashing (De Aceituno et al., 2015), and may not be effective if not prepared properly or if unvalidated recipes are used. Other unsafe practices, including washing produce with soap, have also been promoted online (Geggel, 2020).

The COVID-19 pandemic presents an opportunity to investigate these issues among Canadians. Thus, we conducted an in-depth investigation using qualitative descriptive methods (Neergaard, Olesen, Andersen, & Sondergaard, 2009). The purpose of this study was to investigate how and why Canadians engaged in different food handling and hand hygiene behaviours during the current COVID-19 pandemic. Further, the results of the study can help to identify future public health education, communication, and outreach needs in future times of crisis or emergency.

## Methods

### Study Participants

Online, text-based focus groups were conducted with individuals living in Canada aged 18 or older, who prepared meals at home at least once per week. The focus groups ran from May-June 2020. Participants were recruited via online postings on the Facebook “Focus Groups” group, the Reddit community r/SampleSize, Kijiji.ca (targeting the Canadian cities: Vancouver, Regina, Halifax, Toronto, and Edmonton), and through personal referrals. The recruitment notice directed participants to an online screening form. They were then required to complete this form to ensure they fulfilled eligibility criteria for study inclusion and to provide consent. We contacted eligible participants via a follow-up email to provide details (i.e. date, time, and web-conferencing link) of the focus group. At the end of each focus group, participants were directed to a factsheet detailing best practices for shopping and handling groceries during the pandemic (North Carolina State University, 2020). Each participant was given a $25 (CAD) e-gift card to an online retailer. This study was approved by Ryerson University’s Research Ethics Board (REB #2020-152).

### Data Collection

Participants attended focus groups via the web-conferencing service, Zoom. We used the waiting room and lock-meeting features to ensure meetings were secure. As well, a unique meeting password was required to join each session. Focus groups followed a semi-structured, flexible question guide (Online Resource 1) that was adapted according to the answers provided. The questions were open-ended and inquired about (1) food preparation habits at home, (2) food purchasing, handling, and storage behaviours, (3) hand hygiene and sanitation practices, and (4) information sources about food safety in relation to the COVID-19 pandemic.

We intended to conduct six focus groups consisting of 5-7 participants each, and lasting 1-2 hours in duration (Krueger & Casey, 2015). Two of these focus groups were to include adults aged 40 or older, while the remaining four were to consist of those aged 18-39. The sessions were recorded to ensure that transcripts of the text were available upon completion. The names used in the article are pseudonyms that have been randomly assigned to maintain the confidentiality of participants.

### Data Analysis

Thematic analysis was conducted on the focus group transcripts, using the Theoretical Domains Framework as a coding guide (Atkins et al., 2017). Each individual who participated in a focus group was considered a separate unit of analysis. Once collected, the data were coded by two investigators (I.Y. and R.H.). Both investigators reviewed and coded the first transcript independently, and results were discussed and merged. Minor revisions were also made to the coding form. The remaining six transcripts were then independently coded by both investigators and results were subsequently merged. The coding process entailed numerous read-throughs of the transcripts, identifying trends and patterns, and inferring nuances such as emotion in the data. The analysis was conducted using NVivo12 qualitative analysis software (QSR International, Doncaster, Australia). Themes were developed, revised, refined, and named. Quotes from participants were then selected to best illustrate the main concepts within each theme. Two triangulation methods were utilized to enhance credibility of findings. The use of two data analysts during the coding process provided multiple perspectives on interpretation of the data (Carter, Bryant-Lukosius, DiCenso, Blythe, & Neville, 2014). Member checking was also conducted, which included sending a summary of the results to each participant and asking for their feedback on our interpretation of the overall discussion themes (O’Brien, Harris, Beckman, Reed, & Cook, 2014). The standards for reporting qualitative research (SRQR) guideline was followed during the preparation and reporting of this article (O’Brien et al., 2014).

## Results

Ultimately, seven focus groups were conducted, ranging in size from 2 to 8 participants (n=42 participants total). An additional focus group was added due to low turnout (n=2 participants) in one of the sessions. Participant ages ranged from 19 to 76 years. Four of the focus groups consisted of participants aged 19 – 30 (n = 27), and three of the focus groups consisted of participants aged 40 or older (n = 15). Participants resided in the provinces of British Columbia (n=5), Nova Scotia (n=1), Ontario (n=33), and Saskatchewan (n=3). More participants were female (n=31) than male (n=11). While results showed some age-related differences in behaviours (discussed below), there were not any notable gender differences to report. The member check yielded feedback from one participant, who felt the results were accurately presented.

**Table 1.** Descriptive characteristics of 42 focus group participants, May-June 2020, Canada

| Characteristics | Number of participants (N=42) |
| --- | --- |
| Age (years): |  |
| 18-24 | 8 |
| 25-29 | 9 |
| 30-39 | 10 |
| 40-49 | 10 |
| 50+ | 5 |
| Gender: |  |
| Female | 31 |
| Male | 11 |
| Province of Residence: |  |
| British Columbia | 5 |
| Nova Scotia | 1 |
| Ontario | 33 |
| Saskatchewan | 3 |

Four themes were generated from the coding framework: (1) hand hygiene, grocery shopping, and sanitation practices influenced by the COVID-19 pandemic, (2) the influence of other’s on one’s pandemic experience, (3) emotion and awareness in connection to the pandemic, and (4) routine food safety practices not influenced by the COVID-19 pandemic. The first theme also consists of three subthemes: hand hygiene; grocery shopping; and sanitation and washing of produce. These themes are discussed below, with quotes used to substantiate the main findings within each theme and subtheme.

### Hand hygiene, grocery shopping, and sanitation practices influenced by the COVID-19 pandemic

#### Hand hygiene

Nearly all participants noted improvements in both their frequency and rigour of handwashing. Changes in practices were most pronounced upon returning from outside the home.

> ***Ella:*** *Definitely hand washing more, and very conscious of ‘dirty’ hands. Just feel odd touching anything in the house after going out without purel [sic] or washing with soap and water*.
>
> **Austin:** *Wash my hands more than usual. I’m much more aware of it now*.

Handwashing with soap and water was preferred to using hand sanitizer. Hand sanitizer was primarily used when outside the home (i.e. in the car).

> ***Lana:*** *I only use hand sanitizer when I’m outside the house. When I’m inside I wash if I can. It’s an availability thing as Adrienne said - I prefer washing with soap and water if I can, and would rather avoid public washrooms*

Most people owned hand sanitizer prior to the onset of the pandemic. Several participants noted that quantities were scarce or that the cost had increased, but few reported attempting to make their own.

> ***Ahmed:*** *Had some extra hand sanitizer at home before [the] pandemic. Finally finding a use for them*.
>
> ***Austin:*** *Hard to find!*
>
> ***Jayden:*** *The price is too high now*.

#### Grocery shopping

Most participants reported reducing the frequency of trips to the grocery store. Consequently, they preferred buying higher quantities of foods during these shops to ensure there was enough to last until the next one.

> ***Ahmed:*** *Also didn’t want to go grocery shopping as much as it became a hassle and took much longer. Can’t easily make a quick trip to the grocery store anymore. So bought extras for this reason*.

The types of food people purchased have also changed. Overall, participants purchased more frozen and canned food items, dry goods such as pasta, and, in some cases, pre-packaged produce as participants believed that it would have been handled by fewer people and was therefore “safer” to eat.

> ***Hannah:*** *I bought more canned goods (e.g. beans, corn, etc.), meat to freeze, pasta, frozen foods*.
>
> ***Lana:*** *Not more food in general, but definitely buy more frozen food than before. It’s nice to have stuff that won’t go bad in case you can’t make it to the grocery store etc*.

There were a variety of stockpiling behaviours among participants, ranging from next-to-no stockpiling to families that purchased several months-worth of food supplies. Often, the extent of stockpiling was influenced by available storage space at home.

> ***Grayson:*** *Space in the fridge is of a concern for me and limits how much I can stockpile*.
>
> ***Meera:*** *No as I do not have much space in my fridge/freezer*.

There have been notable changes in shopping behaviours within the grocery store. Participants reported exercising a greater degree of caution (e.g. less face touching, phone use); using gloves, masks, sanitary wipes and/or disposable bags; physical distancing; and browsing more selectively.

> ***Adrienne:*** *When I’m at the grocery store, I feel like everything around me is contaminated and I try to touch as little as I possibly can. I walk around like my hands are covered in mud until I get out of the store and can clean my hands properly (don’t touch my face, don’t touch my phone, etc.)*.

#### Sanitation and washing of produce

Sanitation has been more frequent during the COVID-19 pandemic, with the intensity of people’s rituals varying a great deal. Most participants reported making changes that included sanitizing food packaging and high-touch surfaces with disinfectant wipes. Some established more thorough routines that included step-by-step surface and item sanitation as well as showering and changing clothes upon entering the house. Changes in practices were most pronounced upon returning from outside the home.

> ***Aaliyah****: Non perishables I keep in the car for a couple of days and perishables I bring inside as normal and put away as normal. No washing containers, etc*.
>
> ***Danielle:*** *We use gloves and antiseptic towels*.
>
> ***Angela:*** *At first I would wash my hands first then put on my kitchen gloves before putting groceries away. Didn’t do anything special for the groceries put into the fridge except the mushrooms I would remove from the packaging and put into a paper bag. Then disinfect the handles on the fridge doors and other surfaces I touched. Disinfect the packaging for my dried beans before putting them at away. Then disinfect the cupboard door handles and kitchen faucet, sink*.

For several participants, sanitation practices implemented early in the pandemic have become less strict.

> ***Ella:*** *At first we wiped down all packages with lysol wipes before they entered the house, and a sink full of soapy water to wash all produce before we put it in the fridge. We did this for about a month, and now we don’t do anything*.

There was a significant amount of misinformation reported in terms of washing produce. Participants reported washing their produce with liquid and dish soap, vinegar, and baking soda. Washing with soap was more commonly reported among participants aged 40 and older. Many other participants only washed their produce in water. Of note is that some participants already routinely washed produce with substances in addition to the water before the COVID-19 pandemic.

> ***Lucy:*** *I wash everything down before I put it away. I fill the sink with water and a small dab of liquid soap and wash all fruits and veggies, etc. and then wipe them dry*.

### The influence of others on one’s pandemic experience

Concerns over others’ food handling practices were consistently expressed. This included worries about both grocery store staff and shoppers. Participants were also concerned that grocery store staff might not be taking proper sanitation precautions.

> ***Sarah:*** *I wipe all my items down at the grocery store, and do self check out (because cashiers wear the same gloves for everyone customer) and wash produce before consumption*.
>
> ***Austin:*** *Mostly [worried] from the packaging and who handles it (including the supplier, the grocery store worker, and whoever else might touch it at the store)*.

The reported frequency of purchasing food delivery or takeout has either stayed the same or decreased during the pandemic. Participants’ primary concern surrounding delivery and takeout were the couriers and restaurant staff and their potential to transmit the virus to food and packaging. Further, concerns centred around catching COVID-19, not foodborne illness.

> ***Zoe:*** *I know it might sound a bit much but even when I order grocery or takeout delivery, I think about the person delivering my order and the possibility that they could have it and come into contact with my items*.
>
> ***Julia:*** *Maybe the foods packaging would also concern me. But the courier is the primary*.

Nearly all participants indicated that they sought out information from public health and government officials for COVID-19-related information, primarily through news outlets on television and the internet, as well as government websites. They also believed these sources to be the most trustworthy. However, there were some instances in which medical professionals who were known to the participants had provided inaccurate information or advice that the participant then followed.

> ***Andre:*** *Prime Minister, Premier largely because they are being advised by top medical health professionals*.
>
> ***Noah:*** *I wash [produce] with soap. [Heard this recommendation] through a friend who is [a] Doctor*.
>
> ***Penelope:*** *I like watching the news as it’s interactive; but I also like reading information as I can reread things over*.

There was some discussion over frustration with conflicting or unclear information and how this made participants unsure of what to believe or fostered a sense of distrust.

> **Ahmed:** *I don’t think anyone is an expert at this situation. We may rely on health experts, but clearly they don’t know what’s going on either*.

To some extent, there was an expression of wanting to support others and help keep them safe. This was articulated though desires to support local businesses, forgoing reusable bag use despite their preferences, limiting stockpiling behaviours, and reducing contact with food in grocery stores out of respect for staff and shoppers.

> ***Zoe:*** *I did buy way too much when it was first hitting the news that it was in Ontario. […]. But after that one grocery run, I bought everything in regular amounts (not overdoing it to give others a chance to get what they need)*.

### Emotion and awareness in connection to the pandemic

Several participants indicated a lack of interest or concern over food safety prior to the pandemic but noted that the onset of COVID-19 had alerted them to such concerns.

> ***April****: For me, personally, I’ve never really thought twice about hygiene and my foods. COVID-19 has really changed that for me, I’ve always kind of been a “what doesn’t kill me makes me stronger” type in regard to germs and now I decant boxes as soon as I get home*.

There were a few instances in which shame or embarrassment were expressed in connection to some pandemic-related behaviours.

> ***Hannah:*** *I also (embarrassingly) stockpiled at the beginning of the pandemic*.

Similarly, there was also a sense of unease among some participants when they did not partake in certain behaviours like cleaning food packaging.

> ***Lyla:*** *I think there’s also a sense of panic and guilt if you’re unable to Lysol everything?*
>
> ***Violet:*** *Had concerns about food safety but read somewhere that restaurants practice good hygiene as a rule, so trying to believe that. But handling all the containers is a little anxiety-inducing, to be honest*.

Some participants noted that they were not seeking out information related to food safety during the pandemic. In addition, some participants noted that they were not seeking out any pandemic-related information whatsoever due to concerns over mental health.

> ***Sophie:*** *Haven’t been watching or reading news for that*.
>
> ***Oliver:*** *No reading of that, try to be happy all the time*.

Some participants expressed a desire to continue some of the hygiene and sanitation practices they have developed since the onset of the COVID-19 pandemic, whereas others have indicated that they intend to return to their normal routines as the heightened vigilance and caution causes stress.

> ***Telisha:*** *It definitely has made this concern stronger as in the future, even when restaurants do open up, I think I will be hesitant to dine out before preparing something at home*.
>
> ***Evelyn:*** *I will likely keep up some of the Covid-19 lessons. Living with Sjogren’s makes the common cold a pain. I feel more educated to keep myself healthy from this experience*.
>
> ***Penelope:*** *Once Co-vid is over and there is a vaccine I will return to my old ways. This is way to[o] intense to keep up*.
>
> ***Isabelle:*** *I am not a germaphobe either, I think if I overdo [sic] things it will end up being more stressful*.

### Routine food safety practices not influenced by the COVID-19 pandemic

There appears to be relatively little concern over food safety under “regular” circumstances. Many participants listed vague or non-specific food concerns such as “cross-contamination” or “food poisoning”, and others reported not having food safety concerns whatsoever.

> ***Lyla:*** *I don’t think I had many food safety and security concerns prior to the pandemic*.
>
> ***Sarah:*** *I am not sure that I have one*.
>
> ***Nia:*** *I would have to say cross contamination*.

Almost all participants mentioned utilizing “best before” dates as a means for ensuring their foods were still safe to eat. Participants also heavily relied on their own senses and intuition about the safety of their foods, citing smell, look, texture and sometimes even taste for ways of determining if food was safe to eat.

> ***Julia:*** *I determine its safe to eat by checking the expiration date if it has one. if its fresh produce I look for signs of ripeness versus rotting*.
>
> ***Madison:*** *I smell the product and check the expiry date*.
>
> ***Ella:*** *We’re vegetarian so not often concerned about foods making us sick, unless they have gone mouldy*. ***Keisha****: Sniff test*.

There was a notable range in the length of time that fresh and/or pre-prepared meat and poultry, and leftovers, were kept in the fridge.

> ***Andre:*** *Fresh and ready to eat meat maybe like 3-4 days tops. Leftovers sometimes 1 week at most*.
>
> ***Theodora:*** *[I] never keep raw meat in fridge*.
>
> ***Isaac:*** *We don’t keep leftovers overnight*.

There was very little concern over food storage practices beyond having sufficient space. Most participants did not use refrigerator or freezer thermometers and did not feel they were useful. Again, they relied on their own capabilities and senses to determine whether the appliances were cold enough.

> ***Fatima:*** *Usually, you feel that the fridge/freezer is a certain temp. Obviously, if your fridge stops working you will feel it, so I don’t think it’s necessary to check temp everyday. If my food is going bad I will know its because of the fridge*.

## Discussion

There have been both positive and negative new behaviours adopted by participants since the onset of the COVID- 19 pandemic. The most notable positive change has been the increased frequency and rigour of handwashing. This behaviour change suggests that public health messaging encouraging frequent handwashing that lasts for at least 20 seconds has been communicated and understood by Canadians. Continued efforts are needed to promote the importance of regular handwashing in the future, as proper handwashing is effective in preventing the spread of many communicable diseases in addition to COVID-19 and foodborne illness (Aiello, Coulborn, Perez, & Larson, 2008). Similarly, the additional caution exercised by participants when grocery shopping was a positive change. They appear to be diligent in heeding guidelines related to physical distancing and non-medical mask use.

Conversely, excessive sanitation practices and washing produce in substances other than water alone are two of the recently adopted practices that may result in adverse health consequences. Sanitation practices such as disinfecting grocery packaging, doorknobs, keys, and changing clothes upon returning from outside are likely influenced by results from some early studies, and related media attention, suggesting that virus particles can be recovered from fomites after several days (Rabenau et al., 2005; Van Doremalen et al., 2020). More recent reports, however, suggest that these studies are not reflective of real-life scenarios and that transmission risk from fomites has been exaggerated (Goldman, 2020). Furthermore, of the viral particles recovered in these studies, it is likely that not all are infectious, which may mean that transmission risk is even less pronounced (Lindsley et al., 2010). As such, the extreme effort and caution that many participants reported exercising are unlikely to reciprocate benefits of a similar magnitude. Canadians who engage in such extreme sanitation behaviours may then be inadvertently and unnecessarily increasing their risk of other health issues such as stress and anxiety, as well as accidental poisonings. For example, a 60% increase in calls associated with household cleaner and disinfectant exposures was noted by the British Columbia Centre for Disease Control in mid-March (British Columbia Centre for Disease Control, 2020). Similar spikes in such exposures have also been reported to poison centres across the USA where, during January-March 2020, there was a 20.4% increase in calls compared to January-March 2019 (Chang et al., 2020).

Similarly, the practice of washing produce with soap, vinegar, and/or other substances in addition to water may result in illness due to the ingestion of harmful chemicals or chemical residues (De Pralormo et al., 2019). Some participants reported that they decided to wash their produce in soap by applying the principles of handwashing to produce-washing (i.e. soap breaks down the virus). Others reported that they had heard this suggestion from health professionals. Receiving this type of information from health professionals appears to be common, as seen in an instance in which a family doctor made this recommendation, and others suggesting the importance of extreme sanitation practices for groceries, in a widely-viewed online viral video (Geggel, 2020). It is evident that the lack of explicit information and guidelines surrounding the transmission of the virus from food and packaging, as well as how to handle these items, has played a role in these potentially harmful behaviours. Furthermore, because they are often viewed as reputable sources, it is crucial that health and medical professionals are equipped with accurate knowledge in order to avoid the spread of harmful misinformation.

It should be noted that some participants engaged in the aforementioned produce-washing practices prior to the onset of the COVID-19 pandemic. The apparent lack of understanding and apathy surrounding food safety and associated routine practices not related to the COVID-19 pandemic is cause for concern. For example, our results indicated a heavy reliance on best before dates and “rules of thumb” for determining if frozen and refrigerated foods were safe to eat. However, both results from this study and previous surveys suggest that consumers often do not have a sufficient understanding of best before dates, which apply only to unopened foods (Newsome et al., 2014). This lack of understanding may therefore result in increased instances of foodborne illness. Furthermore, the reported lack of concern and use of refrigerator and freezer thermometers is worrying, as the foodborne pathogen *Listeria monocytogenes* can grow at refrigeration temperatures. This is particularly worrying for older adults, who face an elevated risk of infection, and pregnant women, as listeriosis can result in fetal loss, preterm labour, neonatal sepsis, meningitis, and death (Health Canada, 2016; Silk et al., 2013). Participants in the study also expressed a great deal of apprehension over catching COVID-19 and took precautions against this, yet they had very few concerns over foodborne illness and the potential for infection within the home. These results may indicate a need for improved health promotion campaigns about food safety, especially for high-risk population groups.

In future pandemics or times of crisis, it is essential to have a clear and consistent source to provide information surrounding food safety and sanitation practices, including sanitation of food packaging as well as washing of produce. A trusted, reliable source can help to reduce mixed messages and the resulting food safety behaviours linked to adverse health outcomes (Porter, 2020). Benefits of employing a similar strategy for food safety and sanitation may include both enhancing proper practices that can limit the risk of illness (i.e. due to ingestion of toxic chemicals), as well as reduce unnecessary behaviours (i.e. excessive cleaning) that can result in anxiety, mental fatigue, or accidental poisonings. Our results suggest that this type of messaging would be most well-received when delivered by government and public health officials such as provincial Ministers of Health or Medical Officers of Health, or the Chief Public Health Officer of Canada. As noted in the results, the most effective platforms through which to communicate this information include television and the internet.

There are some limitations to this study. Only four Canadian provinces were represented in this study, and most participants resided in Ontario. Therefore, results may not be representative of all of Canada. Instead, they highlight a range of insights and experiences of Canadians during the COVID-19 pandemic. Additionally, most of the participants were under the age of 40, which likely relates to our online recruitment approach. It would be beneficial to examine these behaviours in more older adults to better compare food handling and hand hygiene behaviours across different age groups. Only one age-related difference was identified in this study (produce-washing practices), which requires further investigation.

Similarly, though both men and women were represented in most of the focus groups, no notable differences in practices were identified. It might be beneficial to conduct future focus groups stratified by gender to help determine if any considerable gender-related differences in behaviours exist. Finally, it is likely that the online, text-based nature of the focus groups resulted in a shorter duration of each focus group. However, the content amassed in online, text-based focus group discussions has been shown to result in comparable quality of data as in-person focus groups (Kite & Phongsavan, 2017; Namey et al., 2020).

## Conclusion

Overall, participants have made several notable changes to their hand hygiene and food handling behaviours during the COVID-19 pandemic. The altered behaviours identified as being the most ubiquitous among participants were handwashing and sanitation practices, including washing of produce in substances other than water alone, as well as exercising greater caution when grocery shopping. Other behaviours, however, such as the stockpiling of perishable foods and the creation of homemade hand sanitizers, were not found to be pervasive. In general, there is a need for better public communication regarding food safety behaviours during the COVID-19 pandemic, and future times of crisis, as concerns around mixed messaging were noted by participants and identified through the reported behaviours. This need for improved communication further extends to routine food safety practices not influenced by the COVID-19 pandemic, as results have highlighted common behaviours that are cause for concern (e.g. use of senses to check when foods are still safe to eat). It is vital to ensure that Canadians have access to trusted and reliable sources of information to inform their food safety and hand hygiene behaviours. Clear, consistent messaging and regular reminders are necessary to reinforce recommended behaviours. Findings from this study can be used to inform future research concerning the effectiveness of food safety and hand hygiene messaging both during and outside of times of crisis.

## Data Availability

The data that support the findings of this study are available from the corresponding author upon reasonable request. The data are not publicly available due to their containing information that could compromise the privacy of research participants.

## Compliance with Ethical Standards

### Ethical Approval

All procedures performed in studies involving human participants were in accordance with the ethical standards of the institution (Ryerson University Research Ethics Board, REB #2020-152) and with the 1964 Helsinki declaration and its later amendments or comparable ethical standards.

### Informed consent

Informed consent was obtained from all individual participants included in the study.

## Author Contributions

Fatih Sekercioglu, Richard Meldrum, and Ian Young contributed to the study conception, design, and material preparation. Data collection and analysis were performed by Robyn Haas and Ian Young. The first draft of the manuscript was written by Robyn Haas, and all authors commented on previous versions of the manuscript. All authors approved the final manuscript.

## References

Aiello, A. E., Coulborn, R. M., Perez, V., & Larson, E. L. (2008). Effect of hand hygiene on infectious disease risk in the community setting: A meta-analysis. American Journal of Public Health, 98(8), 1372–1381. https://doi.org/10.2105/AJPH.2007.124610

Atkins, L., Francis, J., Islam, R., O’Connor, D., Patey, A., Ivers, N., … Michie, S. (2017). A guide to using the Theoretical Domains Framework of behaviour change to investigate implementation problems. Implementation Science, 12(1), 1–18. https://doi.org/10.1186/s13012-017-0605-9

Bialek, S., Boundy, E., Bowen, V., Chow, N., Cohn, A., Dowling, N., … Sauber-Schatz, E. (2020). Severe Outcomes Among Patients with Coronavirus Disease 2019 (COVID-19) — United States, February 12-March 16, 2020. 2020;69:343-346. DOI: http://dx.doi.org/10.15585/mmwr.mm6912e2. *MMWR Morb Mortal Wkly Rep*, 69(12), 343–346. https://doi.org/10.15585/mmwr.mm6912e2

British Columbia Centre for Disease Control. (2020). B.C. sees a spike in calls to poison control about exposure to household cleaners. British Columbia Centre for Disease Control. http://www.bccdc.ca/about/news-stories/stories/2020/b-c-sees-a-spike-in-calls-to-poison-control-about-exposure-to-household-cleaners. Accessed 24 July 2020.

Carter, N., Bryant-Lukosius, D., DiCenso, A., Blythe, J., & Neville, A. J. (2014). The use of triangulation in qualitative research. Oncology Nursing Forum, 41(5), 545–547. https://doi.org/10.1188/14.0NF.545-547

Chang, A., Schnall, A. H., Law, R., Bronstein, A. C., Marraffa, J. M., Spiller, H. A., … Svendsen, E. (2020, April 24). Cleaning and disinfectant chemical exposures and temporal associations with COVID-19 – National poison data system, United States, January 1, 2020- March 31, 2020. Morbidity and Mortality Weekly Report, Vol. 69, pp. 496-498. https://doi.org/10.15585/MMWR.MM6916E1

De Aceituno, A. F., Bartz, F. E., Hodge, D. W., Shumaker, D. J., Grubb, J. E., Arbogast, J. W., … Leon, J. S. (2015). Ability of hand hygiene interventions using alcohol-based hand sanitizers and soap to reduce microbial load on farmworker hands soiled during harvest. Journal of Food Protection, 78(11), 2024–2032. https://doi.org/10.4315/0362-028X.JFP-15-102

De Pralormo, S., Brunet, M., Marquis, A., Bruneau, C., Le Roux, G., & Deguigne, M. (2019). Ingestion of bar soap may produce serious injury: clinical effects and risk factors. Clinical Toxicology, 57(5), 356–361. https://doi.org/10.1080/15563650.2018.1517880

Geggel, L. (2020). Viral video advises washing fruit and vegetables with soap. Here’s why that’s a bad idea. Live Science. https://www.livescience.com/do-not-wash-fruits-vegetables-with-soap.html. Accessed 24 July 2020.

Goldman, E. (2020). Comment Exaggerated risk of transmission of COVID-19 by fomites. The Lancet Infectious Diseases. https://doi.org/10.1016/S1473-3099(20)30561-2

Health Canada. (2016). Risks of listeriosis (Listeria). Government of Canada. https://www.canada.ca/en/public-health/services/diseases/listeriosis/risk-listeriosis.html. Accessed 24 July 2020.

Health Canada. (2019). Safe Food Handling Tips. Government of Canada. https://www.canada.ca/en/health-canada/services/food-nutrition/food-safety/safe-food-handling-tips.html. Accessed 24 July 2020.

Kite, J., & Phongsavan, P. (2017). Insights for conducting real-time focus groups online using a web conferencing service. F1000Research, 6(0), 1–12. https://doi.org/10.12688/f1000research.10427.1

Krueger, R. A., & Casey, M. A. (2015). Focus Groups: A Practical Guide for Applied Research. (5th ed.). SAGE Publications Inc.

Lindsley, W. G., Blachere, F. M., Thewlis, R. E., Vishnu, A., Davis, K. A., Cao, G., … Beezhold, D. H. (2010). Measurements of airborne influenza virus in aerosol particles from human coughs. PloS One, 5(11), e15100. https://doi.org/10.1371/journal.pone.0015100

MacDougall, L., Majowicz, S., Doré, K., Flint, J., Thomas, K., Kovacs, S., & Sockett, P. (2008). Under-reporting of infectious gastrointestinal illness in British Columbia, Canada: Who is counted in provincial communicable disease statistics? Epidemiology and Infection, 136(2), 248-256. https://doi.org/10.1017/S0950268807008461

Murray, R., Glass-Kaastra, S., Gardhouse, C., Marshall, B., Ciampa, N., Franklin, K., … Nesbitt, A. (2017). Canadian consumer food safety practices and knowledge: Foodbook study. Journal of Food Protection, 80(10), 1711–1718. https://doi.org/10.4315/0362-028X.JFP-17-108

Namey, E., Guest, G., O’Regan, A., Godwin, C. L., Taylor, J., & Martinez, A. (2020). How Does Mode of Qualitative Data Collection Affect Data and Cost? Findings from a Quasi-experimental Study. Field Methods, 32(1), 58–74. https://doi.org/10.1177/1525822X19886839

Neergaard, M. A., Olesen, F., Andersen, R. S., & Sondergaard, J. (2009). Qualitative description-the poor cousin of health research? BMC Medical Research Methodology, 9(1), 1–5. https://doi.org/10.1186/1471-2288-9-52

Nesbitt, A., Thomas, M. K., Marshall, B., Snedeker, K., Meleta, K., Watson, B., & Bienefeld, M. (2014). Baseline for consumer food safety knowledge and behaviour in Canada. Food Control, 38(1), 157–173. https://doi.org/10.1016/j.foodcont.2013.10.010

Newsome, R., Balestrini, C. G., Baum, M. D., Corby, J., Fisher, W., Goodburn, K., … Yiannas, F. (2014). Applications and perceptions of date labeling of food. Comprehensive Reviews in Food Science and Food Safety, 13(4), 745–769. https://doi.org/10.1111/1541-4337.12086

North Carolina State University. (2020). COVID-19 and Food Safety FAQ: Shopping and Handling Groceries. North Carolina State University. https://foodsafety.ces.ncsu.edu/wp-content/uploads/2020/04/Handling-Groceries_COVID-19_Flyer-1.pdf?fwd=no. Accessed 24 July 2020.

O’Brien, B. C., Harris, I. B., Beckman, T. J., Reed, D. A., & Cook, D. A. (2014). Standards for reporting qualitative research: A synthesis of recommendations. Academic Medicine, 89(9), 1245–1251. https://doi.org/10.1097/ACM.0000000000000388

Porter, C. (2020). The Top Doctor Who Aced the Coronavirus Test. The New York Times. https://www.nytimes.com/2020/06/05/world/canada/bonnie-henry-british-columbia-coronavirus.html. Accessed 24 July 2020.

Rabenau, H. F., Cinatl, J., Morgenstern, B., Bauer, G., Preiser, W., & Doerr, H. W. (2005). Stability and inactivation of SARS coronavirus. Medical Microbiology and Immunology, 194(1-2), 1-6. https://doi.org/10.1007/s00430-004-0219-0

Silk, B. J., Mahon, B. E., Griffin, P. M., Hannah Gould, L., Tauxe, R. V., Crim, S. M., … Henao, O. L. (2013). Vital signs: Listeria illnesses, deaths, and outbreaks – United States, 2009-2011. Morbidity and Mortality Weekly Report, 62(22), 448–452.

Thomas, M. K., Murray, R., Flockhart, L., Pintar, K., Fazil, A., Nesbitt, A., … Pollari, F. (2015). Estimates of Foodborne Illness-Related Hospitalizations and Deaths in Canada for 30 Specified Pathogens and Unspecified Agents. Foodborne Pathogens and Disease, 12(10), 820–827. https://doi.org/10.1089/fpd.2015.1966

Van Doremalen, N., Bushmaker, T., Morris, D. H., Holbrook, M. G., Gamble, A., Williamson, B. N., … Munster, V. J. (2020, April 16). Aerosol and surface stability of SARS-CoV-2 as compared with SARS-CoV-1. New England Journal of Medicine, Vol. 382, pp. 1564-1567. https://doi.org/10.1056/NEJMc2004973

Wiener-Bronner, D. (2020). Panic buying: How grocery stores restock shelves in the age of coronavirus. CNN. https://www.cnn.com/2020/03/20/business/panic-buying-how-stores-restock-coronavirus/index.html. Accessed 24 July 2020.

Zheng, R., Shou, B., & Yang, J. (2020). Supply disruption management under consumer panic buying and social learning effects. Omega (United Kingdom), 102238. https://doi.org/10.1016/j.omega.2020.102238

